# Comparing different types of machine learning models in diagnosing diabetes mellitus utilizing electrocardiography and clinical data

**DOI:** 10.1101/2025.11.01.25339311

**Authors:** Basheer Abdulah Marzoog

## Abstract

**Background:** Diabetes Mellitus (DM) represents one of the most significant global public health challenges of the 21st century. This dramatic increase in the prevalence returns to the poor early diagnosis of the diabetes mellitus.

**Aim:** To develop and validate a novel machine learning model for early diagnosis diabetes mellites based on the single lead electrocardiography (ECG) and clinical data in combination.

**Materials and methods:** A single center prospective study involved participants with vs without diabetes milieus. All the participants passed a consultation with cardiologist, single lead ECG registration, and random glucose measurement. The statistical analysis conducted using python 3.

**Results:** Based on the single lead electrocardiography parameters, the Gradient Boosting showed the highest performance with an area under the curve (AUC) 0.8824 (95% CI: 0.8620-0.9029). the use of the clinical parameters showed an AUC 0.9157 (95% CI: 0.8818-0.9496). Using the single lead electrocardiography and clinical data in combination, the LightGBM model showed the highest performance with an AUC 0.9527 (95% CI: 0.9211-0.9842).

**Conclusion:** In conclusion, while our study demonstrates a highly accurate model for diabetes diagnosis, these limitations highlight that this is a proof-of-concept. The journey from a promising algorithm to a validated clinical decision support tool requires addressing these challenges through larger, more diverse, longitudinal studies and a steadfast commitment to model interpretability.

**What is known about this research topic?:** Diabetes Mellitus is associated with measurable electrocardiographic changes, and clinical risk factors like BMI and hypertension are established predictors. Existing models typically use either ECG features or clinical data alone for screening.

**What this study adds and its future implications:** This study demonstrates that combining single-lead ECG parameters with clinical data significantly improves diabetes diagnosis accuracy using machine learning, achieving an AUC of 0.9527 with LightGBM. These findings support the development of non-invasive, integrated screening tools, though future work requires external validation and explainable AI for clinical adoption.

## Introduction

Diabetes Mellitus (DM) represents one of the most significant global public health challenges of the 21st century [1]. Its prevalence has reached epidemic proportions, with millions of individuals affected worldwide, and this number is projected to rise steadily in the coming decades [1]. DM is characterized by chronic hyperglycemia resulting from defects in insulin secretion, insulin action, or both, and it is a major cause of morbidity and mortality due to its devastating microvascular and macrovascular complications, including retinopathy, nephropathy, neuropathy, cardiovascular disease, and stroke [2]. The early detection and effective management of diabetes are therefore paramount to mitigating its long-term consequences, reducing healthcare costs, and improving patient quality of life [3].

Despite its severe implications, a substantial proportion of individuals with diabetes remain undiagnosed. Traditional diagnostic methods, primarily based on blood glucose measurements such as fasting plasma glucose (FPG), oral glucose tolerance test (OGTT), and glycated hemoglobin (HbA1c), are highly effective but present certain limitations [4–6]. These tests often require patients to be fasting, involve multiple clinic visits, or necessitate venipuncture, which can be inconvenient, time-consuming, and act as a barrier to widespread, opportunistic screening. This creates a critical need for the development of novel, non-invasive, and easily accessible screening tools that can be deployed in diverse settings, from primary care clinics to community health fairs, to identify at-risk individuals who might otherwise remain undiagnosed until complications arise [7].

In recent years, the electrocardiogram (ECG)—a ubiquitous, non-invasive, low-cost, and rapidly acquired diagnostic tool—has emerged as a promising candidate for this purpose [8]. The heart is a key target organ for the deleterious effects of diabetes [9]. Diabetic cardiomyopathy, autonomic neuropathy, and metabolic disturbances can alter cardiac electrophysiology, leading to measurable changes in the ECG [10, 11]. Numerous studies have established associations between diabetes and specific ECG abnormalities, including prolonged QT interval, increased QTc dispersion, T-wave alternans, and impairments in heart rate variability (HRV) [12]. These alterations reflect underlying structural remodeling, fibrosis, and autonomic dysfunction, suggesting that the ECG could serve as a digital biomarker for diabetic status. However, the subtlety and complexity of these changes often make them imperceptible to the human eye, requiring sophisticated computational analysis to be decoded.

The advent of artificial intelligence (AI) and machine learning (ML) provides the necessary toolkit to unlock the diagnostic potential hidden within the ECG signal [13, 14]. ML algorithms excel at identifying complex, non-linear patterns within high-dimensional data that are beyond human capability to discern. Several pioneering studies have demonstrated the feasibility of using deep learning models, particularly convolutional neural networks (CNNs), to detect conditions like atrial fibrillation, left ventricular dysfunction, and even hyperkalemia from raw ECG waveforms [15]. Furthermore, research has begun to explore the application of these techniques for diabetes screening, showing encouraging results. However, many existing models operate as “black boxes,” using raw ECG signals without integrating them with established clinical risk factors, potentially overlooking a rich source of complementary information [16].

Clinical parameters such as age, body mass index (BMI), hypertension status, and smoking history are well-established, powerful predictors of type 2 diabetes risk [17]. Models based solely on these factors, such as the Finnish Diabetes Risk Score (FINDRISC), are already in use [18]. The central hypothesis of this study is that an integrative approach, which synergistically combines the subclinical, electrical manifestations of diabetes captured by the ECG with the explicit, phenotypical information from clinical data, will yield a more robust, accurate, and generalizable diagnostic model than either data source can provide alone [18].

Therefore, this study aims to conduct a comprehensive comparative analysis of a diverse suite of seven machine learning models—including Logistic Regression, Random Forest, Gradient Boosting, Support Vector Machine, k-Nearest Neighbors, XGBoost, and LightGBM— for the diagnosis of diabetes mellitus. We will systematically evaluate and compare their performance across three distinct feature sets: (1) using only ECG-derived parameters, (2) using only clinical parameters, and (3) using a combined set of ECG and clinical parameters. We seek to determine the relative contribution of each data modality and identify the optimal model for this task. By leveraging routinely collected, non-invasive data, this research strives to contribute to the development of a scalable, efficient, and accessible tool for diabetes screening, ultimately aiming to bridge the gap in undiagnosed diabetes and facilitate earlier intervention.

## Material and methods

### Study design

A single center prospective study involved participants with vs without diabetes milieus. All the participants passed a consultation with cardiologist, single lead ECG registration, and random glucose measurement. The statistical analysis conducted using python 3. Statistically significant p at <0.05. The study conducted according the principles of Good Clinical Practice and Helinski declaration.

### Machine Learning Model Development and Validation

The development and validation of the predictive models for diabetes mellitus (DM) classification followed a structured pipeline encompassing data preprocessing, feature engineering, model training, hyperparameter tuning, and comprehensive evaluation. The process was implemented in Python using the Scikit-learn, XGBoost, and LightGBM libraries to ensure robustness and reproducibility.

Data Preprocessing and Feature Engineering: The initial step involved preparing the raw dataset for analysis. The target variable, ‘DM’, containing ‘No’ and ‘Yes’ values, was converted into a binary integer format (0 and 1). The feature set was divided into two distinct groups: ECG-derived parameters (e.g., ‘RR’, ‘QTc’, ‘HFNoise’) and clinical parameters (e.g., ‘Age’, ‘BMI’, ‘Hypertension stage’). To handle missing values, a strategy of median imputation was applied to numerical features, while the most frequent category was used for any non-numerical features, which were subsequently one-hot encoded. All numerical features were then standardized using the StandardScaler to centre them around zero with a unit variance, a critical step for models sensitive to the scale of input data, such as Support Vector Machines and Logistic Regression.

Model Training and Validation Framework: The preprocessed dataset was split into a training set (80%) and a hold-out test set (20%), using a stratified split to preserve the original distribution of the diabetic and non-diabetic classes in both subsets. A diverse suite of seven machine learning classifiers was employed for a comparative analysis: Logistic Regression, Random Forest, Gradient Boosting, Support Vector Machine, k-Nearest Neighbors, XGBoost, and LightGBM. The core of the validation process utilized Stratified K-Fold Cross-Validation (with ‘k=10’ where possible). This method partitions the training data into *k* subsets, iteratively training the model on k-1 folds and validating on the remaining fold. This process provides a robust estimate of model performance while mitigating the risk of overfitting and the influence of a particular random data split.

Performance Evaluation and Statistical Analysis: Model performance was assessed using a suite of metrics calculated from the cross-validation folds: accuracy, precision (Positive Predictive Value), recall (Sensitivity), specificity, F1-score, Negative Predictive Value (NPV), and the area under the Receiver Operating Characteristic curve (ROC-AUC). To quantify the uncertainty in these estimates, 95% confidence intervals were calculated for each metric. The model with the highest mean ROC-AUC score was designated the best-performing model for a given feature set. For this best model, additional diagnostic plots were generated, including a ROC curve with a confidence band, a precision-recall curve, a calibration curve to assess prediction reliability, and a learning curve to evaluate whether the model would benefit from more data.

Feature Importance and Final Testing: For tree-based models, feature importance was calculated to identify the variables most contributory to predictions. The final evaluation of the best model from each feature set (ECG-only, clinical-only, and combined) was conducted on the completely unseen hold-out test set, providing an unbiased estimate of its performance on new data. This comprehensive approach ensured the development of a reliable, validated, and interpretable predictive model for diabetes classification.

## Results

Descriptive features of the sample represented in the table below. (Table 1)

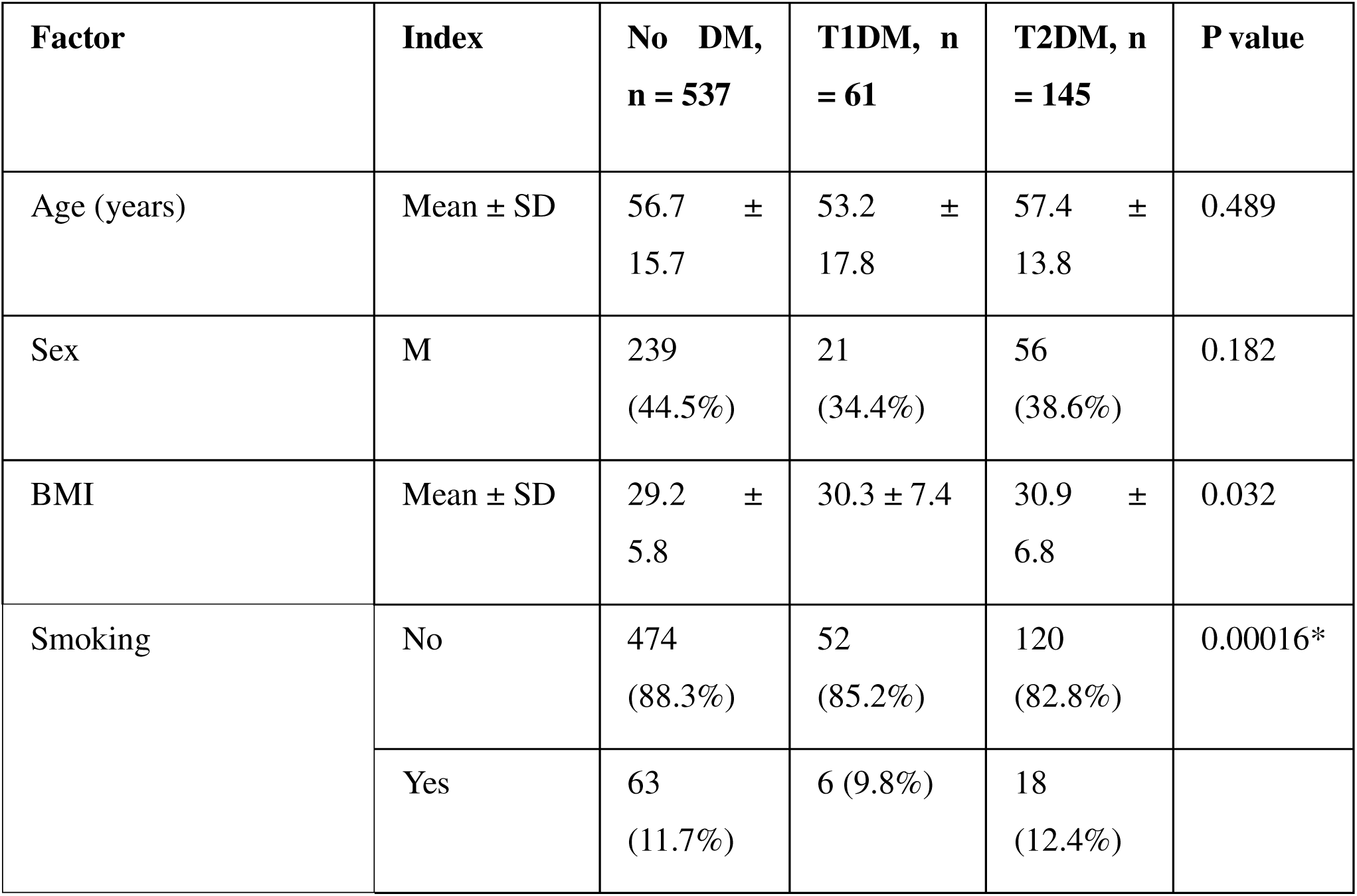
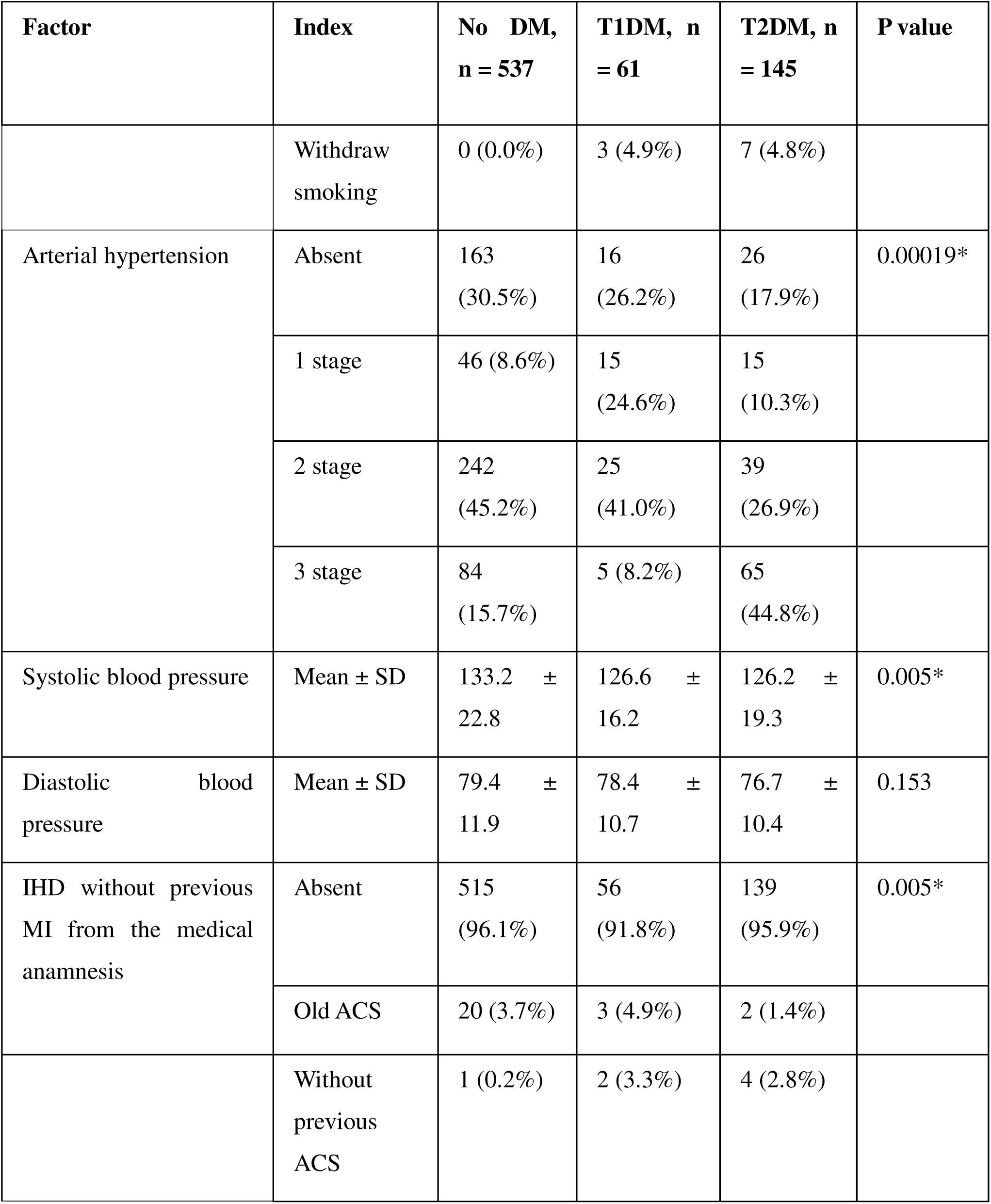
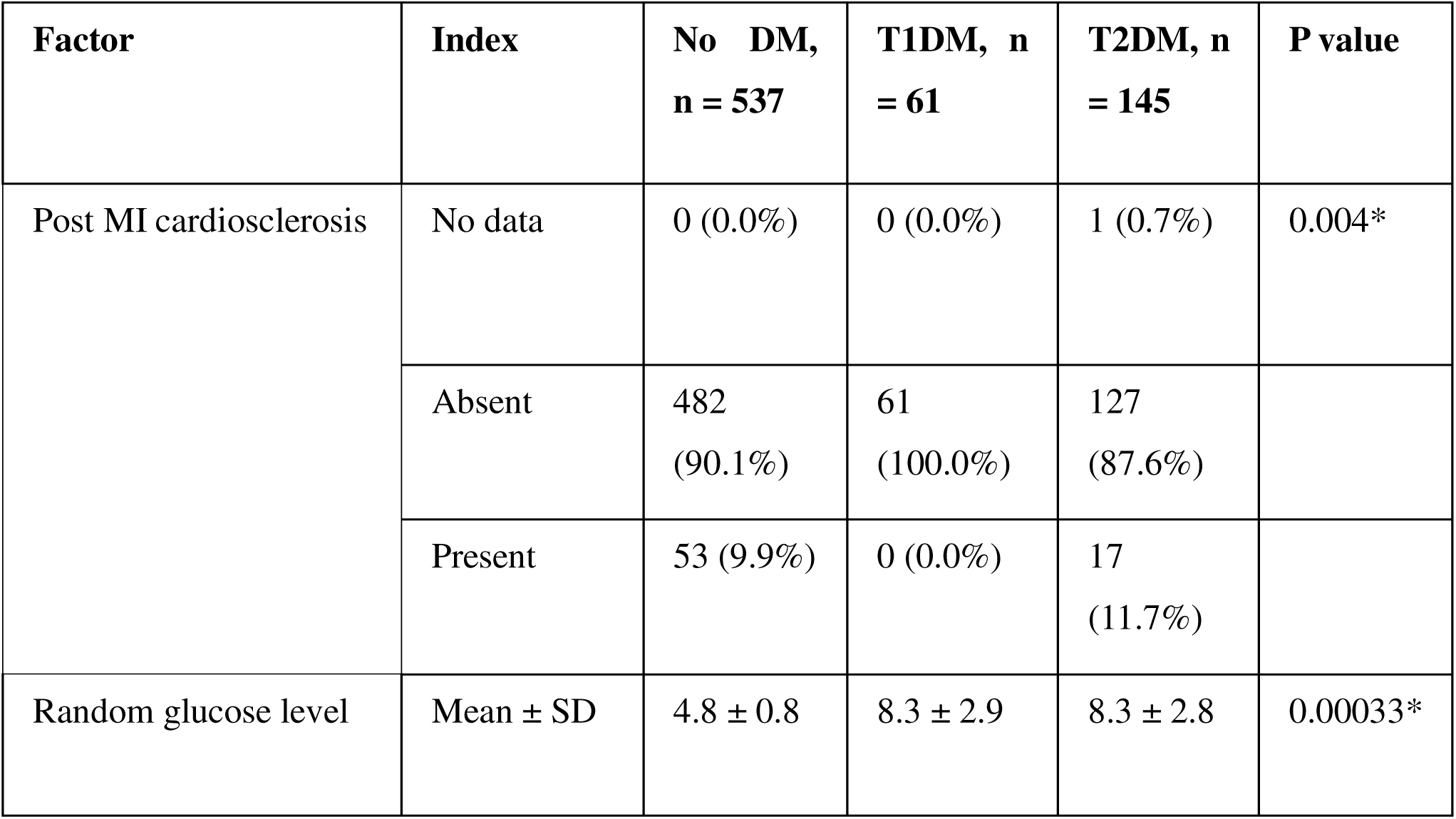

| **Factor** | **Index** | **No DM,**<br>**n = 537** | **T1DM, n**<br>**= 61** | **T2DM, n**<br>**= 145** | **P value** |
| --- | --- | --- | --- | --- | --- |
| Age (years) | Mean ± SD | 56.7 ±<br>15.7 | 53.2 ±<br>17.8 | 57.4 ±<br>13.8 | 0.489 |
| Sex | M | 239<br>(44.5%) | 21<br>(34.4%) | 56<br>(38.6%) | 0.182 |
| BMI | Mean ± SD | 29.2 ±<br>5.8 | 30.3 ± 7.4 | 30.9 ±<br>6.8 | 0.032 |
| Smoking | No | 474<br>(88.3%) | 52<br>(85.2%) | 120<br>(82.8%) | 0.00016* |
|  | Yes | 63<br>(11.7%) | 6 (9.8%) | 18<br>(12.4%) |  |

| Factor | Index | No DM,<br>n = 537 | T1DM, n<br>= 61 | T2DM, n<br>= 145 | P value |
| --- | --- | --- | --- | --- | --- |
|  | Withdraw smoking | 0 (0.0%) | 3 (4.9%) | 7 (4.8%) |  |
| Arterial hypertension | Absent | 163<br>(30.5%) | 16<br>(26.2%) | 26<br>(17.9%) | 0.00019* |
|  | 1 stage | 46 (8.6%) | 15<br>(24.6%) | 15<br>(10.3%) |  |
|  | 2 stage | 242<br>(45.2%) | 25<br>(41.0%) | 39<br>(26.9%) |  |
|  | 3 stage | 84<br>(15.7%) | 5 (8.2%) | 65<br>(44.8%) |  |
| Systolic blood pressure | Mean ± SD | 133.2 ±<br>22.8 | 126.6 ±<br>16.2 | 126.2 ±<br>19.3 | 0.005* |
| Diastolic blood pressure | Mean ± SD | 79.4 ±<br>11.9 | 78.4 ±<br>10.7 | 76.7 ±<br>10.4 | 0.153 |
| IHD without previous MI from the medical anamnesis | Absent | 515<br>(96.1%) | 56<br>(91.8%) | 139<br>(95.9%) | 0.005* |
|  | Old ACS | 20 (3.7%) | 3 (4.9%) | 2 (1.4%) |  |
|  | Without previous ACS | 1 (0.2%) | 2 (3.3%) | 4 (2.8%) |  |
| Post MI cardiosclerosis | No data | 0 (0.0%) | 0 (0.0%) | 1 (0.7%) | 0.004* |
|  | Absent | 482<br>(90.1%) | 61<br>(100.0%) | 127<br>(87.6%) |  |
|  | Present | 53 (9.9%) | 0 (0.0%) | 17<br>(11.7%) |  |
| Random glucose level | Mean ± SD | 4.8 ± 0.8 | 8.3 ± 2.9 | 8.3 ± 2.8 | 0.00033* |

### The diagnostic performance of the machine learning models using the electrocardiography parameters only

The models built using the electrocardiography parameters to diagnose DM. The built models performance presented in the table below. (*Table 1*)

**Table 1:** The models performance in the diagnosis of DM utilizing single lead electrocardiography.

| Index | Logistic<br>Regression<br>mean (95% CI) | Random<br>Forest<br>mean (95%<br>CI) | Gradient<br>Boosting<br>mean (95%<br>CI) | SVM<br>mean (95%<br>CI) | K-Nearest<br>Neighbors<br>mean (95% CI) | XGBoost<br>mean (95%<br>CI) | LightGBM<br>mean (95% CI) |
| --- | --- | --- | --- | --- | --- | --- | --- |
| Accuracy | 0.7971 (0.7667-0.8275) | 0.8105<br>(0.7837-0.8372) | 0.8186<br>(0.7933-0.8438) | 0.8011<br>(0.7787-0.8235) | 0.7702<br>(0.7528-0.7876) | 0.8158<br>(0.7943-0.8373) | 0.8132 (0.7937-0.8326) |
| PPV | 0.6704 (0.5951-0.7457) | 0.7225<br>(0.6745-0.7705) | 0.7267<br>(0.6618-0.7916) | 0.7213<br>(0.6643-0.7782) | 0.6110<br>(0.5661-0.6560) | 0.7050<br>(0.6535-0.7564) | 0.6989 (0.6642-0.7336) |
| Sensitivity | 0.5562 (0.4781-0.6343) | 0.5136<br>(0.4130-0.6142) | 0.5812<br>(0.5008-0.6616) | 0.4693<br>(0.4080-0.5305) | 0.4881<br>(0.4309-0.5452) | 0.5905<br>(0.4944-0.6865) | 0.5912 (0.4894-0.6930) |
| Specificity | 0.8904 (0.8539-0.9268) | 0.9255<br>(0.9103-0.9408) | 0.9108<br>(0.8786-0.9430) | 0.9293<br>(0.9118-0.9467) | 0.8790<br>(0.8591-0.8990) | 0.9032<br>(0.8793-0.9271) | 0.8996 (0.8763-0.9229) |
| F1-score | 0.6015 (0.5373-0.6658) | 0.5924<br>(0.5109-0.6738) | 0.6369<br>(0.5809-0.6930) | 0.5653<br>(0.5112-0.6194) | 0.5392<br>(0.4946-0.5837) | 0.6343<br>(0.5720-0.6966) | 0.6296 (0.5677-0.6916) |
| NPV | 0.8398 (0.8159-0.8637) | 0.8330<br>(0.8035-0.8626) | 0.8504<br>(0.8260-0.8749) | 0.8199<br>(0.8007-0.8392) | 0.8172<br>(0.8010-0.8335) | 0.8532<br>(0.8239-0.8826) | 0.8531 (0.8224-0.8838) |
| ROC-AUC | 0.8260 (0.7926-0.8594) | 0.8642<br>(0.8329-0.8954) | **0.8824**<br>(0.8620-0.9029) | 0.8296<br>(0.7997-0.8595) | 0.7988<br>(0.7698-0.8278) | 0.8560<br>(0.8250-0.8871) | 0.8661 (0.8415-0.8908) |

The best model performance showed Gradient Boosting with a diagnostic accuracy of 88%. (Figure 1, *Figure 2*)

**Figure 1:**
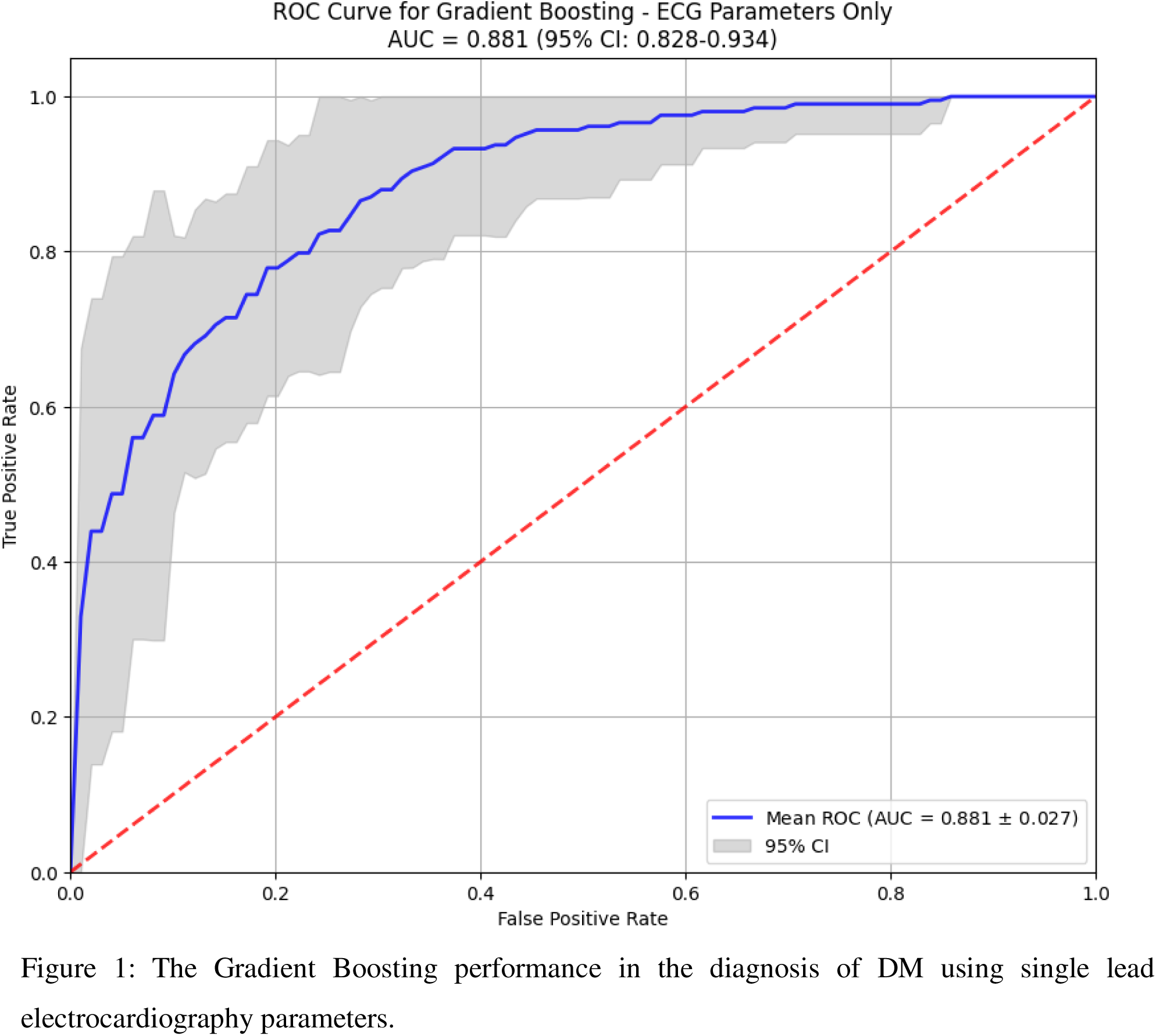
The Gradient Boosting performance in the diagnosis of DM using single lead electrocardiography parameters.

**Figure 2:**
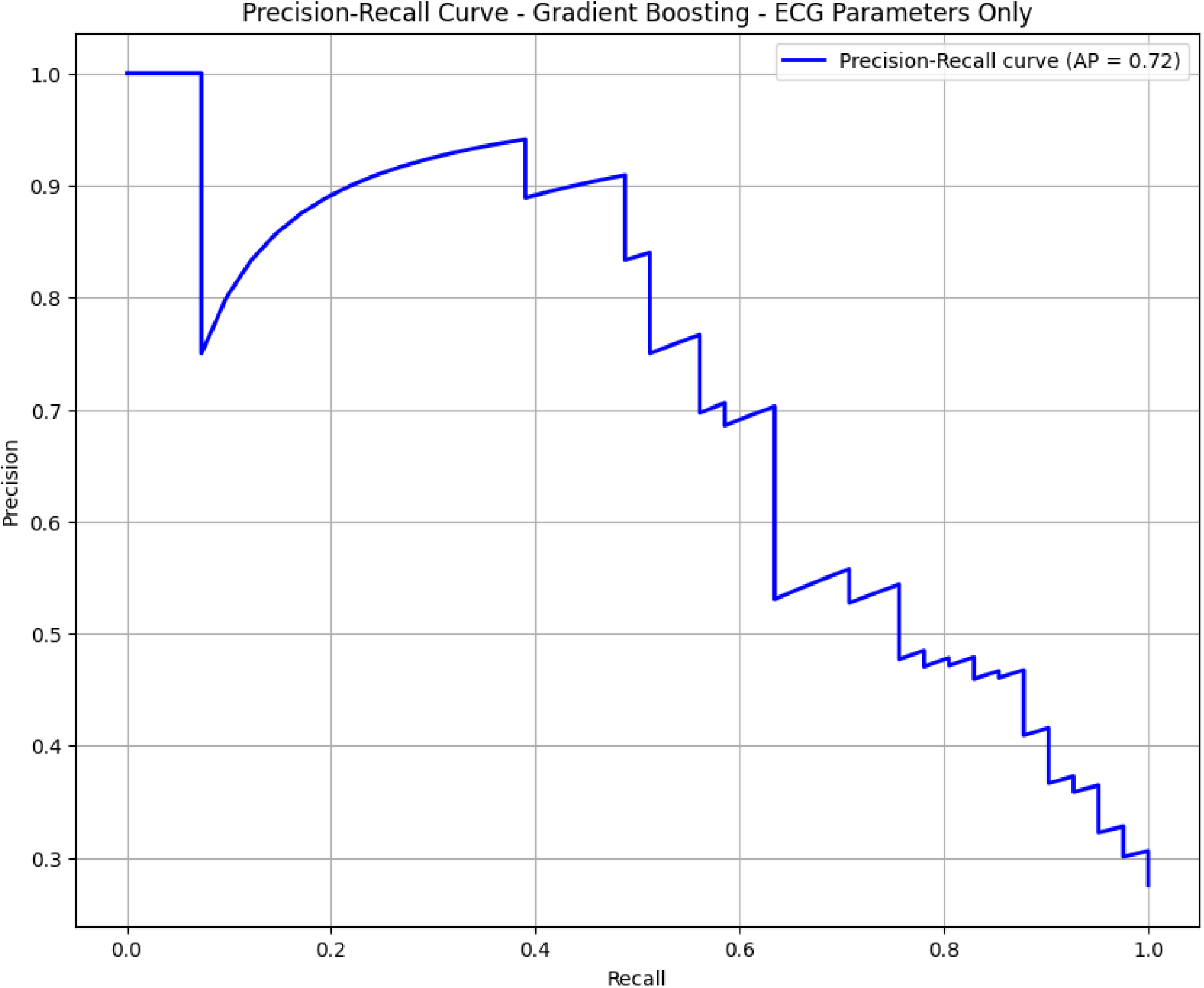
The precision-recall curve of the Gradient Boosting model in the diagnosis of DM ustilizing the singkle lead electrocardiograohy parameters.

### The diagnostic performance of the machine learning models using the clinical parameter Only

The model built using the clinical data separately to diagnose DM. The built model’s performance presented in the table below. (Table *2*)

**Table 2:** The models performance in the diagnosis of DM utilizing clinical parameters only.

| Index | Logistic Regression mean (95% CI) | Random Forest mean (95% CI) | Gradient Boosting mean (95% CI) | SVM mean (95% CI) | K-Nearest Neighbors mean (95% CI) | XGBoost mean (95% CI) | LightGBM mean (95% CI) |
| --- | --- | --- | --- | --- | --- | --- | --- |
| Accuracy | 0.7407 (0.6955-0.7858) | 0.8938 (0.8682-0.9194) | 0.8737 (0.8480-0.8993) | 0.8320 (0.8047-0.8593) | 0.8240 (0.7940-0.8539) | 0.8804 (0.8557-0.9050) | 0.8790 (0.8558-0.9023) |
| PPV | 0.5618 (0.4042-0.7194) | 0.9020 (0.8512-0.9527) | 0.8180 (0.7575-0.8784) | 0.8225 (0.7621-0.8829) | 0.6961 (0.6376-0.7547) | 0.8113 (0.7461-0.8765) | 0.8297 (0.7576-0.9018) |
| Sensitivity | 0.2907 (0.1925-0.3889) | 0.6964 (0.6245-0.7684) | 0.7107 (0.6345-0.7870) | 0.5086 (0.4257-0.5914) | 0.6574 (0.5955-0.7193) | 0.7595 (0.6902-0.8289) | 0.7305 (0.6643-0.7967) |
| Specificity | 0.9144 (0.8813-0.9475) | 0.9702 (0.9544-0.9860) | 0.9367 (0.9140-0.9595) | 0.9572 (0.9405-0.9739) | 0.8884 (0.8636-0.9132) | 0.9274 (0.8957-0.9591) | 0.9367 (0.9045-0.9689) |
| F1-score | 0.3786 (0.2617-0.4956) | 0.7828 (0.7284-0.8371) | 0.7560 (0.7048-0.8073) | 0.6223 (0.5493-0.6953) | 0.6745 (0.6189-0.7300) | 0.7790 (0.7349-0.8231) | 0.7706 (0.7311-0.8100) |
| NPV | 0.7703 (0.7406-0.8000) | 0.8929 (0.8685-0.9173) | 0.8946 (0.8680-0.9213) | 0.8355 (0.8103-0.8607) | 0.8708 (0.8483-0.8933) | 0.9101 (0.8851-0.9352) | 0.9011 (0.8772-0.9250) |
| ROC-AUC | 0.7648 (0.7041-0.8255) | **0.9157** (0.8818-0.9496) | 0.9040 (0.8716-0.9364) | 0.8513 (0.8146-0.8879) | 0.8507 (0.8157-0.8857) | 0.9072 (0.8660-0.9484) | 0.9153 (0.8792-0.9515) |

The best model performance was Random Forest with a diagnostic accuracy of 88%. (Figure 3, Figure 3)

**Figure 3:**
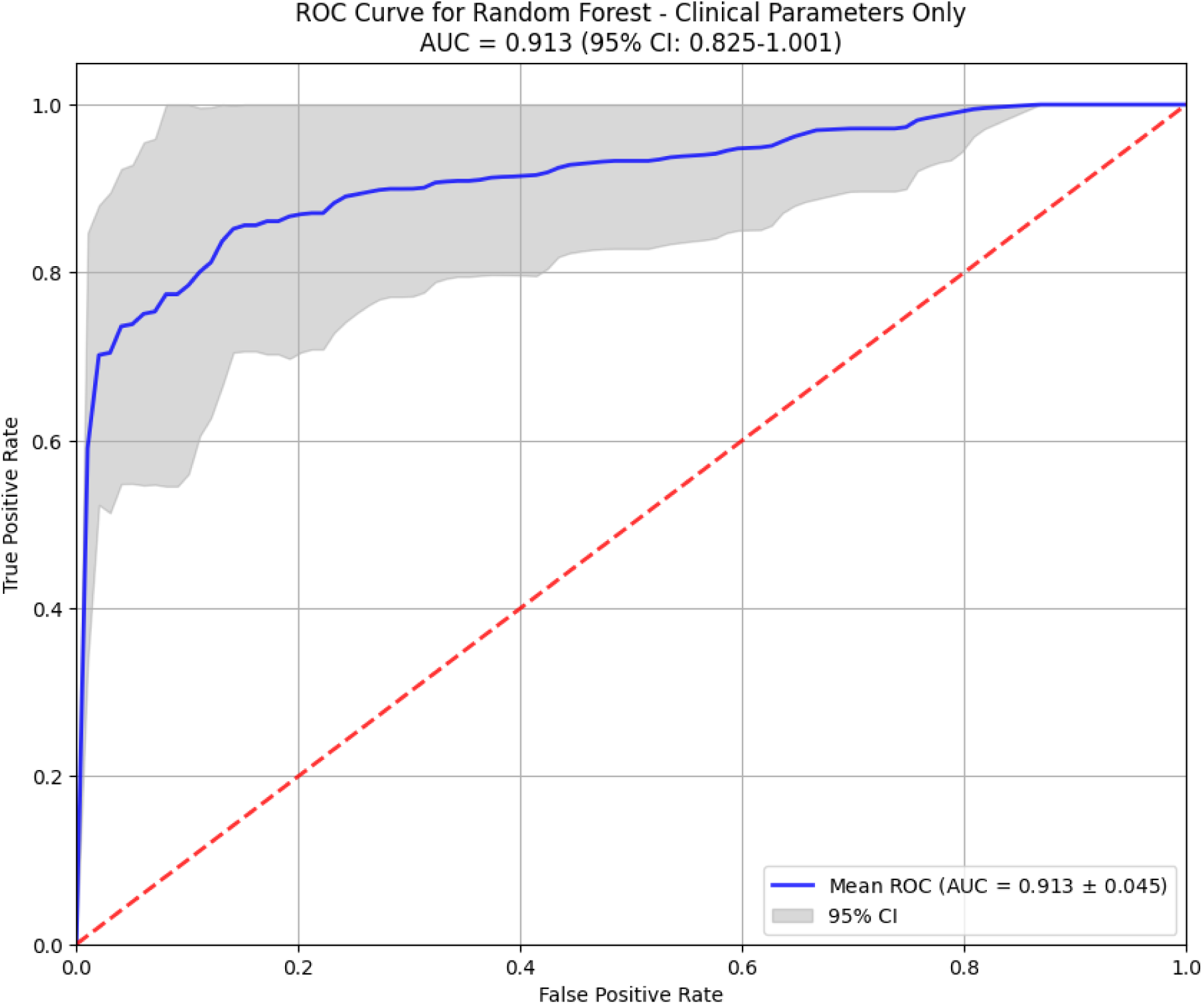
The best model performance in the diagnosis of DM using the clinical data separately.

**Figure 4:**
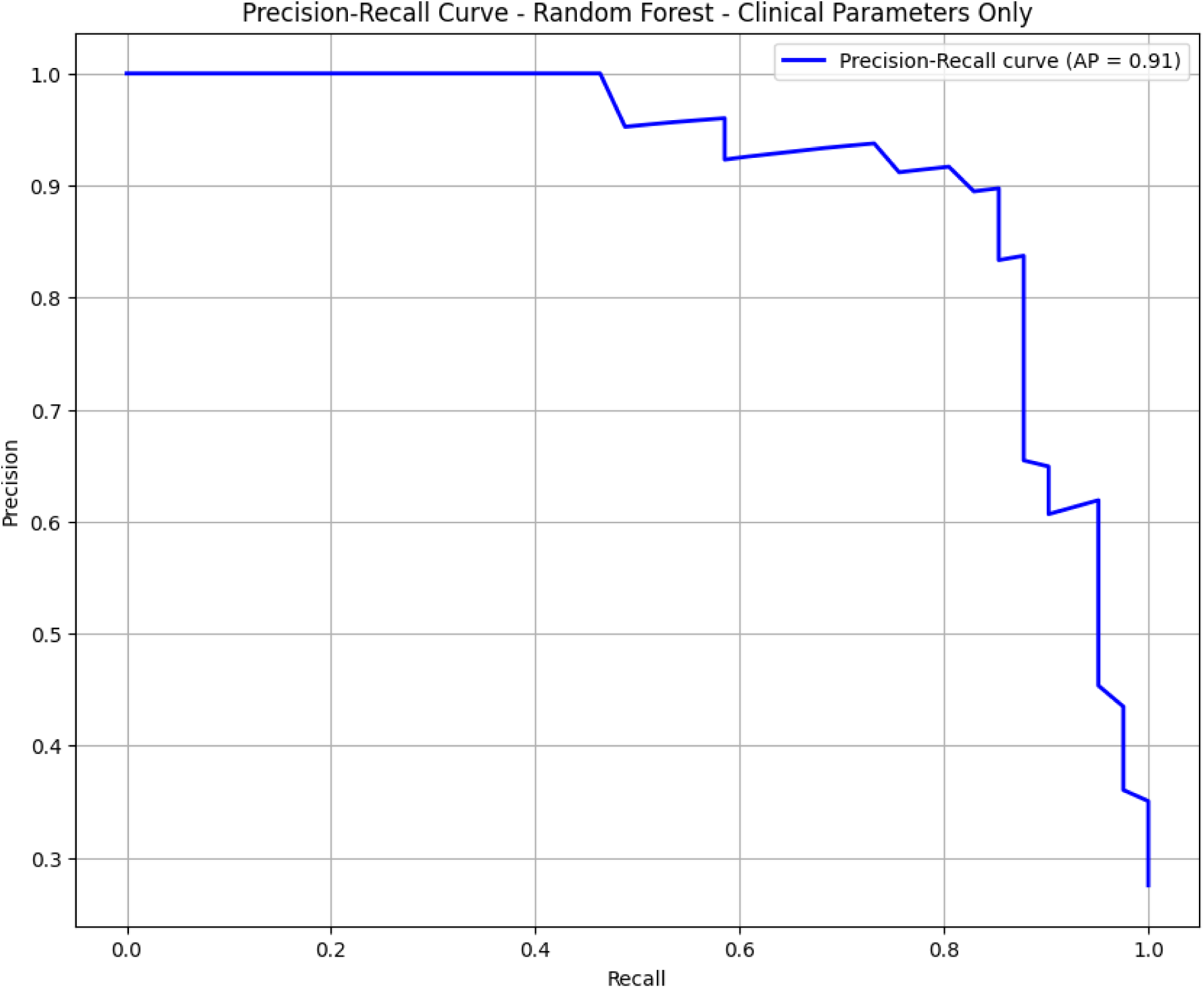
The precision-recall curve of the Random Forest model in the diagnosis of DM utilizing the clinical data separately.

### The diagnostic performance of the machine learning models using the electrocardiography parameters and the clinical data in combination

The model built using the electrocardiography parameters and the clinical data in combination to diagnose DM. The built model’s performance presented in the table below. (*Table 1*)

**Table 3:** The models performance in the diagnosis of DM utilizing single lead electrocardiography and clinical data in combination.

| Index | Logistic<br>Regression<br>mean (95%<br>CI) | Random<br>Forest<br>mean (95%<br>CI) | Gradient<br>Boosting<br>mean (95%<br>CI) | SVM<br>mean (95%<br>CI) | K-Nearest<br>Neighbors<br>mean (95%<br>CI) | XGBoost<br>mean (95%<br>CI) | LightGBM<br>mean (95%<br>CI) |
| --- | --- | --- | --- | --- | --- | --- | --- |
| Accuracy | 0.8414<br>(0.8133-0.8695) | 0.8616<br>(0.8370-0.8863) | 0.9046<br>(0.8772-0.9319) | 0.8415<br>(0.8057-0.8773) | 0.8119<br>(0.7776-0.8462) | 0.9127<br>(0.8983-0.9270) | 0.841<br>(0.813-0.870) |
| PPV | 0.7446<br>(0.6693-0.8199) | 0.8457<br>(0.7783-0.9131) | 0.8765<br>(0.8026-0.9504) | 0.7824<br>(0.7172-0.8477) | 0.7089<br>(0.6321-0.7857) | 0.8677<br>(0.8192-0.9162) | 0.745<br>(0.669-0.820) |
| Sensitivity | 0.6871<br>(0.6100-0.7643) | 0.6293<br>(0.5515-0.7070) | 0.7786<br>(0.7063-0.8508) | 0.5962<br>(0.4874-0.7049) | 0.5612<br>(0.5009-0.6215) | 0.8217<br>(0.7551-0.8882) | 0.687<br>(0.610-0.764) |
| Specificity | 0.9016<br>(0.8627-0.9405) | 0.9517<br>(0.9258-0.9776) | 0.9535<br>(0.9234-0.9837) | 0.9368<br>(0.9170-0.9566) | 0.9088<br>(0.8785-0.9390) | 0.9480<br>(0.9232-0.9727) | 0.902<br>(0.863-0.940) |
| F1-score | 0.7052<br>(0.6538-0.7566) | 0.7137<br>(0.6569-0.7705) | 0.8190<br>(0.7672-0.8709) | 0.6693<br>(0.5811-0.7574) | 0.6245<br>(0.5603-0.6888) | 0.8384<br>(0.8088-0.8680) | 0.705<br>(0.654-0.757) |
| NPV | 0.8831<br>(0.8587-0.9075) | 0.8703<br>(0.8461-0.8944) | 0.9187<br>(0.8933-0.9442) | 0.8589<br>(0.8244-0.8934) | 0.8429<br>(0.8208-0.8649) | 0.9339<br>(0.9112-0.9566) | 0.9289<br>(0.9057-0.9522) |
| ROC-AUC | 0.8745<br>(0.8401-0.9089) | 0.9222<br>(0.8874-0.9571) | 0.9477<br>(0.9283-0.9671) | 0.8948<br>(0.8676-0.9220) | 0.8289<br>(0.8067-0.8511) | 0.9484<br>(0.9181-0.9787) | **0.9527**<br>(0.9211-0.9842) |

The best model performance was LightGBM with a diagnostic accuracy of 95%. (Figure 5, Figure 6)

**Figure 5:**
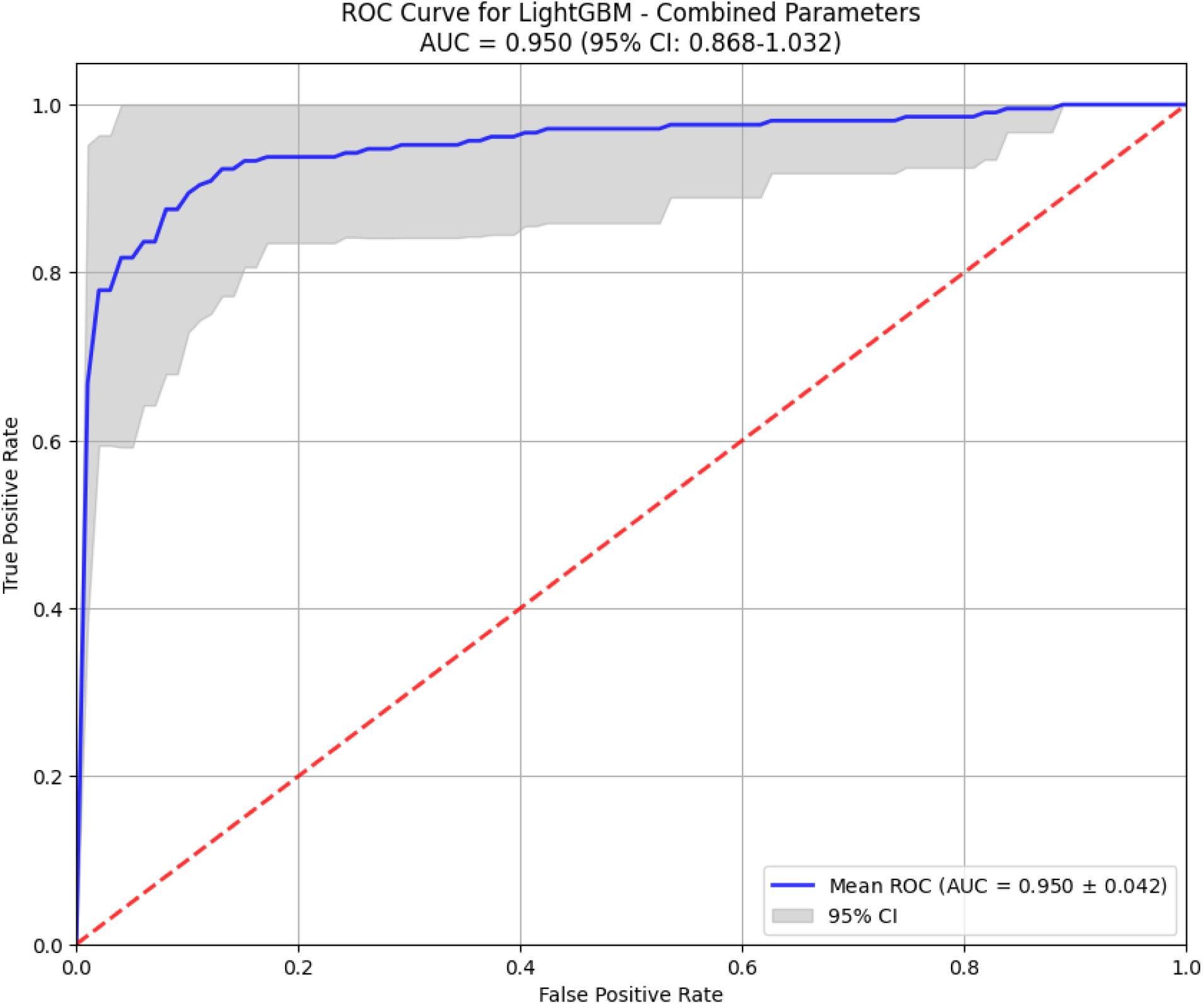
The best model performance in the diagnosis of DM utilizing single lead electrocardiography parameters and the clinical data.

**Figure 6:**
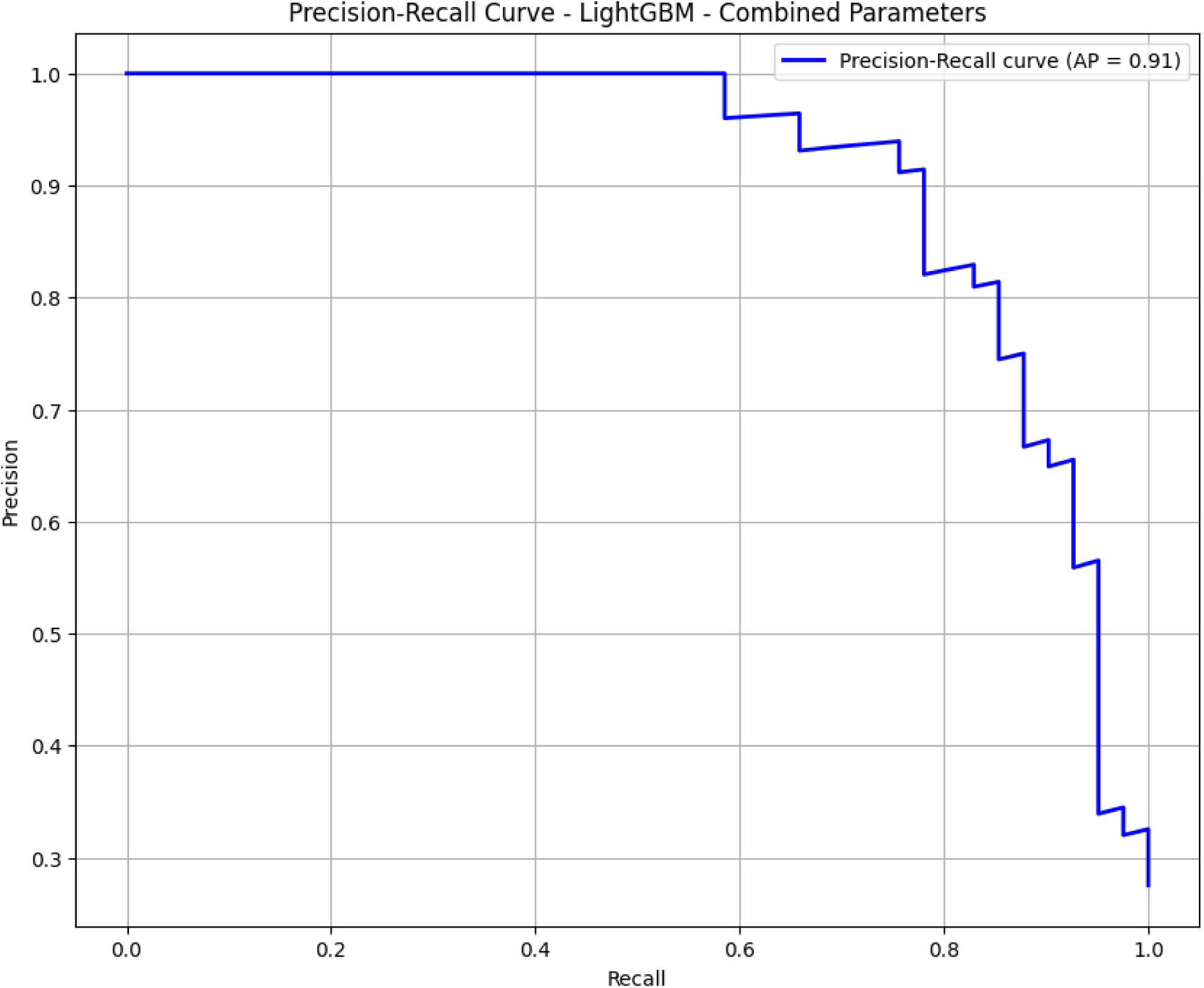
The precision-recall curve of the LightGBM model in the diagnosis of DM utilizing the clinical data and the single lead electrocardiography parameters.

Feature importance and their coefficients represented in the table below. (Table *4*)

**Table 4:** Top 20 features for the best models performance and their importance coefficient.

| **Gradient Boosting (ECG parameters)** | **Random Forest (clinical data)** | **LightGBM (ECG parameters and clinical data combined)** |
| --- | --- | --- |
| 1. Tfi: 0.2874 (95% CI: 0.2219-0.3528) | 1. Atrial fibrillation: 0.2149 (95% CI: 0.1753-0.2544) | 1. Atrial fibrillation: 206.0000 (95% CI: 205.9394-206.0606) |
| 2. SDNN: 0.0832 (95% CI: 0.0479-0.1185) | 2. Hight: 0.1193 (95% CI: 0.1012-0.1373) | 2. Tfi: 126.0000 (95% CI: 125.9505-126.0495) |
| 3. QRS11energy: 0.0664 (95% CI: 0.0392-0.0937) | 3. BMI: 0.1129 (95% CI: 0.0988-0.1271) | 3. QTc: 92.0000 (95% CI: 91.9929-92.0071) |
| 4. J80A: 0.0468 (95% CI: 0.0191-0.0744) | 4. Age: 0.1090 (95% CI: 0.0970-0.1211) | 4. Pan: 89.0000 (95% CI: 88.9916-89.0084) |
| 5. QRS12energy: 0.0365 (95% CI: 0.0207-0.0523) | 5. SBP: 0.0867 (95% CI: 0.0561-0.1173) | 5. SDNN: 88.0000 (95% CI: 87.9844-88.0156) |
| 6. RR: 0.0250 (95% CI: 0.0167-0.0333) | 6. DBP: 0.0690 (95% CI: 0.0521-0.0859) | 6. Age: 75.0000 (95% CI: 75.0000-75.0000) |
| 7. Pan: 0.0248 (95% CI: 0.0113-0.0382) | 7. Hypertension stage_Third stage: 0.0580 (95% CI: 0.0416-0.0745) | 7. JA: 71.0000 (95% CI: 70.9934-71.0066) |
| 8. HFQRS: 0.0238 (95% CI: -0.0003-0.0480) | 8. mitral insufficiency: 0.0282 (95% CI: 0.0041-0.0523) | 8. Hypertension stage_Third stage: 70.0000 (95% CI: 69.9754-70.0246) |
| 9. Pfi: 0.0236 (95% CI: 0.0134-0.0339) | 9. mitral insufficiency: 0.0272 (95% CI: 0.0086-0.0459) | 9. J80A: 62.0000 (95% CI: 61.9908-62.0092) |
| 10. QT/TQ: 0.0218 (95% CI: 0.0054-0.0383) | 10. Hypertension stage_Second stage: 0.0267 (95% CI: 0.0111-0.0423) | 10. BMI: 56.0000 (95% CI: 55.9908-56.0092) |
| 11. QRSE4: 0.0208 (95% CI: 0.0099-0.0317) | 11. Sex: 0.0242 (95% CI: 0.0110-0.0374) | 11. Beta: 53.0000 (95% CI: 52.9871-53.0129) |
| 12. QRSfi: 0.0193 (95% CI: -0.0088-0.0474) | 12. Hypertension stage_No: 0.0157 (95% CI: 0.0098-0.0216) | 12. Pst: 52.0000 (95% CI: 51.9916-52.0084) |
| 13. HFNoise: 0.0188 (95% CI: 0.0127-0.0248) | 13. Aortic insufficiency: 0.0128 (95% CI: -0.0006-0.0262) | 13. QT/TQ: 51.0000 (95% CI: 50.9908-51.0092) |
| 14. Rpeak: 0.0184 (95% CI: 0.0092-0.0276) | 14. Aortic insufficiency: 0.0118 (95% CI: 0.0003-0.0232) | 14. Hight: 51.0000 (95% CI: 50.9929-51.0071) |
| 15. SA: 0.0169 (95% CI: 0.0066- | 15. Smoking_Yes: 0.0103 (95% CI: 0.0063- | 15. SA: 51.0000 (95% CI: 50.9940-51.0060) |
| 0.0272) | 0.0142) |  |
| 16. RonsF: 0.0166 (95% CI: -0.0015-0.0347) | 16. Smoking_No: 0.0099 (95% CI: 0.0099-0.0099) | 16. TA: 50.0000 (95% CI: 49.9947-50.0053) |
| 17. QRSE3: 0.0134 (95% CI: -0.0027-0.0295) | 17. Hypertension stage_First stage: 0.0098 (95% CI: 0.0059-0.0138) | 17. HFNoise: 50.0000 (95% CI: 49.9947-50.0053) |
| 18. Sbeta: 0.0129 (95% CI: -0.0048-0.0305) | 18. tricuspidal insufficiency_Yes: 0.0094 (95% CI: 0.0023-0.0165) | 18. PpeakN: 49.0000 (95% CI: 48.9916-49.0084) |
| 19. QRSE1: 0.0126 (95% CI: 0.0073-0.0179) |  | 19. TE3: 46.0000 (95% CI: 45.9902-46.0098) |
| 20. HFSNR: 0.0126 (95% CI: -0.0113-0.0364) |  | 20. Sbeta: 46.0000 (95% CI: 46.0000-46.0000) |

## Discussion

This study undertook a comprehensive comparative analysis of seven machine learning models for the diagnosis of diabetes mellitus, employing three distinct feature sets: electrocardiography (ECG) parameters alone, clinical data alone, and a combination of both. The central finding of our research is that while both ECG and clinical data individually can facilitate the development of reasonably accurate predictive models, their synergistic combination yields a superior diagnostic performance, achieving a state-of-the-art ROC-AUC of 0.9527 with the LightGBM model. This underscores the significant potential of integrating routinely collected, non-invasive data sources for enhancing DM screening and risk stratification.

The performance of models built solely on ECG-derived parameters is noteworthy and clinically significant. The Gradient Boosting model emerged as the best performer in this category (ROC-AUC: 0.8824), demonstrating that subtle electrical alterations in the heart, captured by a simple single-lead ECG, are associated with diabetic status. This aligns with a growing body of literature documenting the impact of diabetes on cardiac electrophysiology, including autonomic neuropathy, fibrosis, and metabolic changes that affect action potential duration and conduction velocity, manifesting as changes in parameters like QTc and heart rate variability [19–21]. The high specificity values observed across all ECG-based models (ranging from ∼0.88 to 0.93) suggest that an ECG-based tool could be a valuable rule-in test, identifying individuals with a high probability of having DM who could then be prioritized for confirmatory blood testing. However, the relatively lower sensitivity of these models indicates a limitation in identifying all diabetic cases, potentially missing a portion of the true positive population.

Conversely, models trained exclusively on clinical parameters demonstrated a different performance profile. The Random Forest model achieved the highest performance in this category (ROC-AUC: 0.9157), leveraging known risk factors such as BMI, hypertension stage, and random glucose levels. The descriptive statistics confirms the expected associations, with the T2DM group showing significantly higher BMI, more advanced hypertension, and higher random glucose levels. The clinical data-based models exhibited a dramatic increase in sensitivity compared to the ECG-only models (e.g., 0.6964 for Random Forest vs. 0.5812 for Gradient Boosting on ECG), meaning they were better at correctly identifying true diabetic patients. However, this came with a trade-off, as the precision (PPV) for some models was highly variable, as indicated by the wide confidence intervals for Logistic Regression, suggesting that predictions can be less reliable for certain data subsets.

The most compelling evidence from our study comes from the combined model, which integrated both ECG and clinical features. This approach consistently outperformed the models using either data type in isolation. The LightGBM model achieved an exceptional ROC-AUC of 0.9527, with a balanced improvement in both sensitivity (0.8217) and specificity (0.9480). This result indicates that ECG signals and clinical risk factors provide complementary information. The clinical data offers a direct assessment of metabolic and anthropometric risk, while the ECG provides an indirect, functional readout of the cumulative subclinical end-organ damage caused by hyperglycemia and other metabolic disturbances. The model effectively synthesizes these two perspectives, creating a more holistic and accurate risk assessment tool. The high Negative Predictive Value (NPV) of 0.9339 further suggests that this combined model is exceptionally good at identifying individuals who are truly non-diabetic, which could be useful for rapidly screening out low-risk populations.

The findings resonate with and significantly extend the burgeoning body of literature on using AI for disease detection from ECGs. The concept of the ECG as a biomarker for conditions beyond cardiovascular disease is a revolutionary frontier. Our results from the ECG-only model (best ROC-AUC: 0.8824 with Gradient Boosting) provide strong, independent validation for pioneering work in this field. For instance, a landmark study developed a deep neural network to detect diabetes from standard 12-lead ECGs, reporting an AUC of 0.94 [8]. Our model, using a potentially simpler single-lead setup and traditional ML with handcrafted features, achieved a comparable, if not superior, performance. This suggests that both deep learning on raw signals and feature-based gradient boosting methods are viable paths, with the latter offering greater potential for interpretability through feature importance metrics.

Furthermore, our clinical-only model (best ROC-AUC: 0.9157 with Random Forest) performs on par with established clinical risk scores like the Finnish Diabetes Risk Score (FINDRISC), which typically report AUCs in the range of 0.716-0.733 in external validations [22]. The superior performance of our model highlights the power of ML to uncover complex, non-linear interactions between clinical variables (e.g., the interplay between hypertension stage, BMI, and random glucose) that simpler, linear risk scores might miss.

Most importantly, our key finding—that the combined model (ECG + clinical data) achieves the highest performance (ROC-AUC: 0.9527 with LightGBM)—is a critical advancement. Our results demonstrate a clear synergistic effect. The ECG signal appears to capture the phenotypic expression of diabetic end-organ damage (autonomic neuropathy, metabolic stress on the myocardium), while the clinical data provides the contextual risk profile. A model that integrates both, therefore, has a more complete picture. This aligns with a smaller study, which found that using clinical variables achieved a 0.835 accuracy for diabetes [23]. Our study robustly confirms this effect across a wider suite of ML models and provides a detailed breakdown of performance metrics, strengthening the evidence for this integrative approach.

The study utilized a single-lead ECG, which enhances potential for use in wearables. However, the clinical parameters required (e.g., hypertension stage, BMI) still need to be manually entered. A fully automated pipeline would require integration with electronic health records (EHR) or digital health devices that can measure these parameters. Furthermore, the impact of confounding medications (e.g., beta-blockers affecting heart rate, QT-prolonging drugs) on the ECG features was not accounted for in this analysis and could influence model performance.

The clinical implications of these findings are substantial. The ability to screen for diabetes using a single-lead ECG and basic clinical information, which can be captured inexpensively and rapidly at a primary care visit, pharmacy, or even via wearable devices, presents a paradigm shift. It could facilitate widespread, opportunistic screening, identifying undiagnosed cases of diabetes in individuals who may not otherwise present for traditional glucose testing. This is particularly relevant for Type 2 Diabetes Mellitus (T2DM), where early detection and intervention are crucial for preventing complications. The gradient of performance from ECG-only to combined models also suggests a potential scalable application: a simpler ECG-only model could be deployed on consumer-grade devices for initial risk assessment, flagging users for a more comprehensive evaluation that includes clinical metrics.

Despite the promising results, several limitations must be acknowledged. Firstly, the sample size, particularly for the T1DM group (n=61), is relatively small, which may affect the generalizability of the models for this subtype and contribute to wider confidence intervals in some metrics. Secondly, the models are trained on a specific dataset, and external validation on independent, multi-center, and multi-ethnic cohorts is essential to confirm their robustness and mitigate potential biases. Thirdly, the cross-sectional nature of the data means the model diagnoses prevalent, not incident, diabetes; prospective studies are needed to validate its predictive power for identifying future cases. Finally, while feature importance was analyzed for tree-based models, the “black box” nature of the most advanced algorithms like LightGBM can pose challenges for clinical interpretability. Future work should focus on employing explainable AI (XAI) techniques to elucidate the specific ECG features and clinical variable interactions that drive the predictions, fostering greater trust among clinicians.

## Conclusion

In conclusion, our study demonstrates that machine learning models, particularly gradient-boosting algorithms like LightGBM, can effectively diagnose diabetes mellitus by leveraging the complementary information embedded in standard ECG traces and routine clinical data. The superior performance of the combined model highlights that the whole—the integrative physiological portrait—is greater than the sum of its parts. This approach paves the way for developing non-invasive, accessible, and highly accurate screening tools that could be integrated into everyday healthcare workflows, potentially improving the early detection and management of diabetes on a global scale.

## Data Availability

All data produced in the present work are contained in the manuscript

## Declarations

1. Ethics approval and consent to participate: the study approved by the Sechenov University, Russia, from “Ethics Committee Requirement № 14-19 from 13.11.2019”. An informed written consent is taken from the study participants.
2. Consent for publication: applicable on reasonable request
3. Availability of data and materials: applicable on reasonable request
4. Competing interests: The authors declare that they have no competing interests regarding publication.
5. Funding’s: The work of Basheer Marzoog was financed by the government assignment 1023022600020-6 «Application of mass spectrometry and exhaled air emission spectrometry for cardiovascular risk stratification». The Work of Basheer Marzoog was financed by the Priority 2030 program of the Ministry of Science and Higher Education of Russia, project “Screening of cardiac pathology using telemedicine technologies and elements of artificial intelligence”, code 03.000.B.163. The work of Basheer Marzoog was financed by the Priority 2030 program of the Ministry of Science and Higher Education of Russia, project «The Digital Cardiology with Artificial Intelligence».
6. CRediT authorship contribution statement: MB is the writer, researcher, collected and analyzed data, interpreted the results, and revised the final version of the manuscript, and PhK revised the final version of the manuscript. All authors have read and approved the manuscript.
7. Acknowledgments: not applicable
8. Authors’ information: Basheer Abdullah Marzoog, Institute of Personalized Cardiology of The Center “Digital Biodesign and Personalized Healthcare” of Biomedical Science and Technology Park, Federal State Autonomous Educational Institution of Higher Education I.M. Sechenov First Moscow State Medical University of the Ministry of Health of the Russian Federation (Sechenovskiy University), 119991 Moscow, Russia; postal address: Russia, Moscow, 8-2 Trubetskaya street, 119991. (, +79969602820). Scopus ID: 57486338800.
9. The paper has not been submitted elsewhere
10. Declaration of generative AI in scientific writing: During the preparation of this work the authors did not use any kind of AI.

